# Factors associated with accelerated resting heart rate among college freshmen : a cross-sectional study

**DOI:** 10.1101/2020.07.31.20165969

**Authors:** Qiqi You, Tiantian Wang, Qingqing Jiang, Qiumei Zhang, Shiyi Cao

**Affiliations:** School of Public Health, Tongji Medical College, Huazhong University of Science and Technology, Wuhan 430030, Hubei, China; Hospital of Jianghan University, Wuhan 430056, Hubei, China

**Keywords:** resting heart rate, freshman, college, prevalence

## Abstract

**Objective:** This study aimed to investigate the prevalence of accelerated resting heart rates (RHRs) among freshmen in a university in Wuhan and to explore the influencing factors.

**Methods:** We conducted a cross-sectional survey and physical examinations in a university in Wuhan from 2015 to 2017, including 12428 freshmen. A binary logistic regression model was undertaken to identify the influencing factors associated with their accelerated RHRs.

**Results:** The prevalence of accelerated RHRs was 47.5% among the 12428 freshmen surveyed. Accelerated RHR of freshmen was related to female (odds ratio [OR]: 1.484, 95% confidence interval [95% CI]: 1.368 - 1.610, *P* < 0.001) and high blood pressure (OR: 2.709, 95% CI: 2.212 - 3.318, *P* < 0.001). Compared with rural students, accelerated RHR was more likely to occur in urban students (OR: 1.452, 95% CI: 1.333 - 1.583, *P* < 0.001). Additionally, students who came from the central and the eastern regions (OR: 1.452, 95% CI: 1.254 - 1.682, *P* < 0.001; OR: 1.363, 95% CI: 1.149 - 1.618, *P* < 0.001), rather than who came from the western regions, were more likely to have accelerated RHRs.

**Conclusions:** Students with accelerated RHRs made up a high proportion of college freshmen entering the university in 2015 – 2017 in Wuhan. For people aged around 18 years, more attention was needed to pay to RHRs and specific measures such as blood pressure management were required in advance to prevent accelerated RHRs.

**Key questions:** *What is already known about this subject?:* Accelerated resting heart rate (RHR) could significantly increase the risk of suffering from cardiovascular diseases (CVDs). However, little research had reported the influencing factors associated with accelerated RHR among the young around 18 years old.

*What does this study add?:* We conducted a survey on freshmen to investigate the influencing factors associated with accelerated RHR, so as to provide early warning information for the occurrence of CVDs in a visual way.

*How might this impact on clinical practice?:* This study might serve as a reminder to young people to pay more attention to RHRs and provide early warning information about CVDs.

## Introduction

Resting heart rate (RHR), as a easily measurable indicator, was the number of beats per minute of the heart in an awake and quiet environment, which now was commonly used to reflect the excitability of the sympathetic nerves^1^. In a previous study, RHR over 80 beats per minute was called accelerated RHR^2^. Although accelerated RHR had not yet been defined as a disease, it made adverse effects on people’s health, and the impact might be more serious than tachycardia. Because its effects were subtle, and people could not pay enough attention to it.

Studies had shown that accelerated RHR might increase the risk of suffering from atherosclerosis^3, 4^ and cardiovascular diseases (CVDs)^5-8^. The World Health Organization had reported that CVDs were the leading cause of death globally: more people died annually from CVDs than from any other cause. And an estimated 17.9 million people died from CVDs in 2016, representing 31% of all global deaths. The acceleration of RHR might result in uneven recovery of cardio-myocytes, unstable ventricular electromyography activities and lower threshold of ventricular fibrillation, which made rapid arrhythmia, sudden cardiac death and other adverse cardiovascular events more likely to occur. At the same time, RHR could reflect the balance between autonomic nerves^1^. When accelerated RHR appeared, the autonomic nervous system was already in an unbalanced state, which might cause lipid metabolism disorder, inflammation^9, 10^ and other damages to the body.

Many studies on RHR had been carried out at home and abroad, mainly researching on the association between CVDs and RHR^5-8^. However, studies on the factors affecting RHR were limited. A few studies^11-13^ had explored the influencing factors of RHR, but only in middle-aged and elderly people. Although CVDs were usually manifested in the elderly, the process that could not be observed obviously might occur at a younger age^14, 15^. In recent years, there was an increasing tendency of mortality of CVDs among the young^16^. Therefore, the detection and prevention of CVDs in young people were extremely urgent. Hence, we conducted a survey on freshmen in a university in Wuhan to find out the prevalence of RHR of people aged around 18 years and to investigate its influencing factors, so as to provide early warning information for the occurrence of CVDs in a visual way.

## Methods

### Ethics statement

All participants were informed of the purpose of this investigation and signed informed consent forms. The investigation was approved by the Research Ethics Committee in Tongji Medical College, Hua Zhong University of Science and Technology, Wuhan, China.

### Settings and study subjects

The cross-sectional study was an investigation of the prevalence of accelerated RHR among college freshmen and the influencing factors associated with it. We conducted this investigation from 2015 to 2017 in a university in Wuhan. Inclusion criteria of participants: (1) They did not suffer from arrhythmia or any other disease that affected the heart rate; (2) They did not use drugs that might influence the heart rate. Finally, we identified 12428 college freshmen to take part in this survey.

### Data collection

On the basis of literature research and expert consultation, standard-structured questionnaires were designed and then distributed to the participants. Students who met the criteria had orderly physical examinations organized by the college. The questionnaires and physical examination forms were recycled. The contents of the questionnaire were as follows: department, major, enrollment year, age, gender, residence, region, “exercise (at least 150 minutes of physical activities per week) or not”, “sleep late (sleep duration ≤ 6 hours/day) or not”, and “have a family history of cardiovascular diseases (CVDs) (first - degree relatives have a history of CVDs) or not”. Physical measurements included height, weight, blood pressure, RHR and so on. Height and weight were measured with Height and Weight Meter. The measurement methods of blood pressure and RHR referred to the *National Grassroots Hypertension Prevention and Management Guidelines*.

### Explanatory Variables

Based on the formula for calculating body mass index (BMI), we calculated the value of BMI of each participant and classified them according to the diagnostic criteria for overweight and obesity. BMI between 18.5 kg/m^2^ and 23.9 kg/m^2^ was classified as “normal weight”, BMI between 24 kg/m^2^ and 27.9 kg/m^2^ was classified as “overweight”, and BMI equal or greater than 28 kg/m^2^ was classified as “obesity”; Classification standards for minors^17^ referred to Table 3. According to the diagnostic criteria in *Chinese Guidelines for The Prevention and Treatment of Hypertension*^18^, we divided systolic blood pressure (SBP) < 120 mmHg and diastolic blood pressure (DBP) < 80 mmHg as “normal blood pressure”, 120 mmHg ≤ SBP < 140 mmHg and/or 80 mmHg ≤ DBP < 90 mmHg as “normal high blood pressure”, and SBP ≥ 140 mmHg and/or DBP ≥ 90 mmHg as “high blood pressure”.

### Statistical analysis

Data from questionnaires and physical examinations were double-entry by Epidata 3.0 and statistical analyses were performed using SPSS 22. Pearson chi-square tests were used to compare the prevalence of accelerated RHR between groups. Binary logistic regression analysis was performed to determine the factors associated with accelerated RHRs among college freshmen. All comparisons were two-tailed, and *P*-values < 0.05 were considered statistically significant.

## Results

In summary, 12428 students were enrolled in the study, 56% of whom were women. Among them, 4406 students were enrolled in 2015, 4020 students were enrolled in 2016, and 4002 students were enrolled in 2017. The mean age of the participants was 18.4 (standard deviation (SD) = 0.8) years. Overall, 5900 students (47.5%) had accelerated RHRs.

Table 1 shows the frequency of accelerated RHRs in each group according to different characteristics of college freshmen and shows the results of intergroup comparisons. Pearson chi-square tests indicated that RHR was associated with students’ age, gender, residence, region, BMI, blood pressure and whether students regularly exercised (*P* < 0.05). Notably, compared with college freshmen with normal RHRs, students who aged less than 18 years, who were females, who lived in urban areas, who came from the central and the eastern regions, who were obese, whose blood pressure were high, and who did not regularly exercise were more likely to have accelerated RHRs. However, no significant differences were observed between the two groups in whether students slept late (*P* = 0.305) and whether students had a family history of CVDs (*P* = 0.078).

**Table 1.**
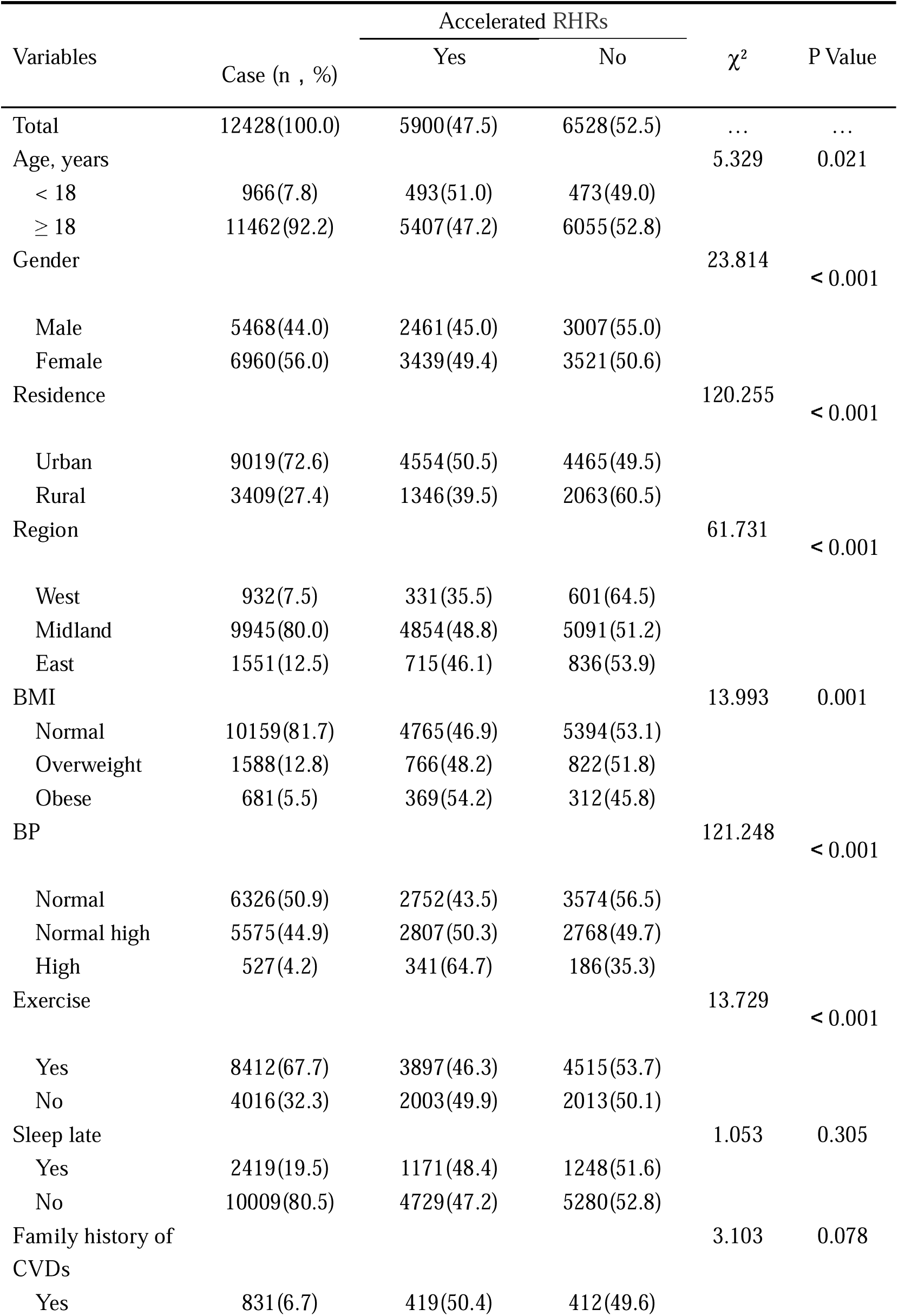
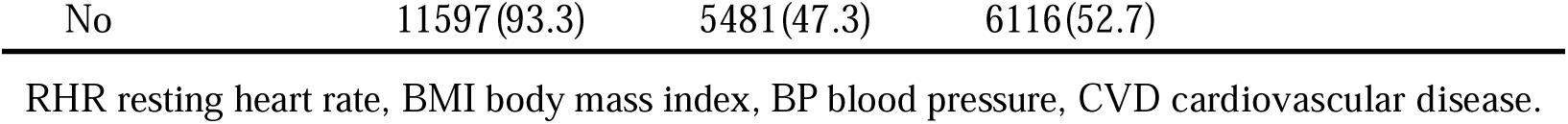
Characteristics of College Freshmen and Associations with Accelerated RHRs.

| Variables | Case (n , %) | Accelerated RHRs | | $\chi^2$ | P Value |
| --- | --- | --- | --- | --- | --- |
|  |  | Yes | No |  |  |
| Total | 12428(100.0) | 5900(47.5) | 6528(52.5) | ... | ... |
| Age, years |  |  |  | 5.329 | 0.021 |
| < 18 | 966(7.8) | 493(51.0) | 473(49.0) |  |  |
| ≥ 18 | 11462(92.2) | 5407(47.2) | 6055(52.8) |  |  |
| Gender |  |  |  | 23.814 | < 0.001 |
| Male | 5468(44.0) | 2461(45.0) | 3007(55.0) |  |  |
| Female | 6960(56.0) | 3439(49.4) | 3521(50.6) |  |  |
| Residence |  |  |  | 120.255 | < 0.001 |
| Urban | 9019(72.6) | 4554(50.5) | 4465(49.5) |  |  |
| Rural | 3409(27.4) | 1346(39.5) | 2063(60.5) |  |  |
| Region |  |  |  | 61.731 | < 0.001 |
| West | 932(7.5) | 331(35.5) | 601(64.5) |  |  |
| Midland | 9945(80.0) | 4854(48.8) | 5091(51.2) |  |  |
| East | 1551(12.5) | 715(46.1) | 836(53.9) |  |  |
| BMI |  |  |  | 13.993 | 0.001 |
| Normal | 10159(81.7) | 4765(46.9) | 5394(53.1) |  |  |
| Overweight | 1588(12.8) | 766(48.2) | 822(51.8) |  |  |
| Obese | 681(5.5) | 369(54.2) | 312(45.8) |  |  |
| BP |  |  |  | 121.248 | < 0.001 |
| Normal | 6326(50.9) | 2752(43.5) | 3574(56.5) |  |  |
| Normal high | 5575(44.9) | 2807(50.3) | 2768(49.7) |  |  |
| High | 527(4.2) | 341(64.7) | 186(35.3) |  |  |
| Exercise |  |  |  | 13.729 | < 0.001 |
| Yes | 8412(67.7) | 3897(46.3) | 4515(53.7) |  |  |
| No | 4016(32.3) | 2003(49.9) | 2013(50.1) |  |  |
| Sleep late |  |  |  | 1.053 | 0.305 |
| Yes | 2419(19.5) | 1171(48.4) | 1248(51.6) |  |  |
| No | 10009(80.5) | 4729(47.2) | 5280(52.8) |  |  |
| Family history of CVDs |  |  |  | 3.103 | 0.078 |
| Yes | 831(6.7) | 419(50.4) | 412(49.6) |  |  |

|  |  |  |  |
| --- | --- | --- | --- |
| No | 11597(93.3) | 5481(47.3) | 6116(52.7) |

RHR resting heart rate, BMI body mass index, BP blood pressure, CVD cardiovascular disease.

Table 2 presents the factors associated with accelerated RHRs by binary logistic regression model. Factors significantly associated with students’ accelerated RHRs included: female (odds ratio [OR]: 1.484, 95% confidence interval [95% CI]: 1.368 - 1.610, *P* < 0.001), living in urban areas (OR: 1.452, 95% CI: 1.333 - 1.583, *P* < 0.001), coming from the central (OR: 1.452, 95% CI: 1.254 - 1.682, *P* < 0.001) and the eastern regions (OR: 1.363, 95% CI: 1.149 - 1.618, *P* < 0.001), and high blood pressure (OR: 2.709, 95% CI: 2.212 - 3.318, *P* < 0.001). There were no significant differences in the risk of accelerated RHRs between groups that whether students regularly exercised (*P* = 0.471), whether students slept late (*P* = 0.743), and whether students had a family history of CVDs (*P* = 0.526). In addition, no significant difference was observed in the risk of accelerated RHRs among freshmen in different BMI groups.

**Table 2.** Factors Associated with Accelerated RHRs of College Freshmen in Binary Logistic Regression Model.

| Variables | OR | 95% CI | P Value |
| --- | --- | --- | --- |
| Age ≥ 18 years (Ref = age < 18 years) | 0.872 | 0.763-0.997 | 0.045 |
| Female (Ref = male) | 1.484 | 1.368-1.610 | <0.001 |
| Urban (Ref = rural) | 1.452 | 1.333-1.583 | <0.001 |
| Region (Ref = west) |  |  |  |
| Midland | 1.452 | 1.254-1.682 | <0.001 |
| East | 1.363 | 1.149-1.618 | <0.001 |
| BMI (Ref = normal) |  |  |  |
| Overweight | 0.946 | 0.843-1.062 | 0.347 |
| Obese | 1.087 | 0.916-1.289 | 0.341 |
| BP (Ref = normal) |  |  |  |
| Normal high | 1.456 | 1.333-1.590 | <0.001 |
| High | 2.709 | 2.212-3.318 | <0.001 |
| Exercise (Ref = not exercise) | 0.966 | 0.880-1.061 | 0.471 |
| Sleep late (Ref = not sleep late) | 1.016 | 0.923-1.118 | 0.743 |
| Family history of CVDs (Ref = not have a family history of CVDs) | 0.954 | 0.824-1.104 | 0.526 |

OR odds ratio, CI confidence interval, BMI body mass index, BP blood pressure, CVD cardiovascular disease.

**Table 3.** Guidelines for the prevention and control of overweight and obesity in school-age children in China.

| Age, years | Overweight (BMI, kg/m<sup>2</sup>) |  | Obesity (BMI, kg/m<sup>2</sup>) |  |
| --- | --- | --- | --- | --- |
|  | male | female | male | female |
| 15 | 23.1 | 23.4 | 26.9 | 26.9 |
| 16 | 23.5 | 23.7 | 27.4 | 27.4 |
| 17 | 23.8 | 23.8 | 27.8 | 27.7 |

BMI body mass index.

## Discussion

This study showed that the prevalence of accelerated RHRs of freshmen in the university from 2015 to 2017 was 47.5%. And the mean value of RHR was 80.3 (SD = 10.0) beats/min (79.6 ± 10.3 beats/min for males and 80.8 ± 9.7 beats/min for females), which was a little higher than adolescents in Brazil in Pedro C Hallal’ research (78.4 beats/min)^19^. Compared with the middle-aged and the elderly in other researches, the average RHR of college freshmen was also higher^20^, indicating that RHRs in people aged around 18 years tended to be faster.

Results showed that living in urban areas, as well as coming from the central and the eastern regions, were associated with accelerated RHRs, mainly due to lifestyle differences. Compared with rural and western areas, urban and eastern areas were more developed and the pace of life was more intense, hence, people living in urban areas or coming from the eastern regions were under more pressure, as were students. A previous study showed^21^ that psychological stress could act as a “stressor”, prompting human body to produce stress responses, leading to excessive activation of sympathetic system, and then promoting the release of catecholamines, finally accelerating the RHR.

Consistent with previous studies, we found that accelerated RHR was associated with high blood pressure^11, 22^. The effect between high blood pressure and RHR was bidirectional. At present, many studies reported that accelerated RHR could increase the risk of high blood pressure^23-26^. On the one hand, accelerated RHR affected blood pressure by increasing cardiac contraction; on the other hand, accelerated RHR made vascular endothelial dysfunction, leading to a decrease in the release of various vasodilatory substances, while an increase in the release of vasoconstrictor substances, which eventually elevated the blood pressure. Conversely, high blood pressure also made impact on RHR. Studies had shown that patients with high blood pressure were usually accompanied by accelerated RHRs. And as the blood pressure increased, the incidence of accelerated RHR gradually increased. Heart rate was an important indicator of sympathetic excitability. Sympathetic excitability was higher in people with high blood pressure than in people with normal blood pressure. In addition, heart rate fluctuations, heart rate variability, norepinephrine levels in plasma, and muscular sympathetic excitability were all higher in hypertensive patients than in non-hypertensive patients, which might influence RHR^27, 28^.

Contrary to previous reports, our analysis showed no association between accelerated RHR and BMI or exercise. In our study, no significant association was found between obesity and accelerated RHR. However, Wang F^22^ found that the relative risk of obesity resulting in accelerated RHR was 0.530. Obesity was not a risk factor of RHR but a protective factor. Therefore, whether obesity accelerated RHR was controversial and more research was needed. In addition, previous studies^29, 30^ had also shown that regular exercise could slow down people’s RHRs, especially in athletes, related to autonomic tone changing or sinus node remodeling. However, we found that whether students exercised had no significant effect on RHRs. The difference in participants might explain the source of difference in this result, and more similar studies were needed to verify the specific result.

The strengths of the current study were as follows. Firstly, the included participants were about 18 years, who were easily exposed to disease-related risk factors but did not have adequate disease prevention awareness. This investigation highlighted the early prevention of accelerated RHR, and played an early warning role in the occurrence of CVDs. Secondly, the sample size of this study was relatively large, and the results were relatively reliable. This study had also a few limitations. First of all, this study was a cross-sectional study, which could only provide clues to the etiology, and further exploration was needed to prove the causal relationship; Secondly, accelerated RHRs of some students might be caused by other non-specific factors, such as mood, breathing and posture changes, which cannot be avoided.

## Data Availability

The data came from the standard-structured questionnaire surveys and physical examinations of freshmen in a university in Wuhan from 2015 to 2017.

## Authors’ Contributions

QZ contributed to the conception or design of the work. QJ contributed to the acquisition or interpretation of data for the work. QY and TW contributed to the analysis of data and drafted the manuscript. SC contributed to guiding article writing. All authors critically revised the manuscript and gave final approval of the article.

## Acknowledgements

We thank all college freshmen involved in the study, as well as teachers in the university in Wuhan for their help.

## Declaration of Conflicting interests

The authors declare that there is no conflict of interest.

## Funding

This work was supported by the Fundamental Research Funds for the Central Universities, Huazhong University of Science and Technology [grant number 2020WKZDJC015].

## Notes

### Competing Interest Statement

The authors have declared no competing interest.

### Author Declarations

The investigation was approved by the Research Ethics Committee in Tongji Medical College, Hua Zhong University of Science and Technology, Wuhan, China.

